# Violence and aggression towards Doctors in Pakistan: a nationwide survey

**DOI:** 10.1101/2025.07.11.25331344

**Authors:** Muhammad Daniyal, Abdul Latif, Sardar Noman Qayyum, Sadia Tameez-ud-din, Taaha Ahmad, Tehreem Fatima, Naeem khattak, Syed Niaz Hussain Shah, Safi Ullah, Miraj Ahmad, Zar Khan

## Abstract

**Introduction:** Although workplace violence impacts nearly all sectors, it is especially prevalent in healthcare settings, raising serious concerns for public health and occupational safety. Workers in healthcare and social services face the highest rates of violence-related injuries, being five times more likely to be injured compared to those in other industries. Furthermore, incidents of violence in the health sector are estimated to represent approximately 25% of all workplace violence occurrences

**Objective:** The main aim of this study is to examine the frequency of exposure of doctors to violence and aggression at work, and the contributing factors thereon, as well as the effects of violence on doctors.

**Method:** This study has been conducted in multiple hospitals in Pakistan’s five provinces (Punjab, Sindh, Khyber Pakhtunkhwa (KPK), Baluchistan, and Gilgit Baltistan), the Capital (Islamabad), and Azad Jammu and Kashmir (AJK). The questionnaire prepared by the researchers was filled in through face-to-face meetings by 1003 healthcare professionals who agreed to take part in the study. The resulting data has been assessed and evaluated using the SPSS V27 package program.

**Results:** 77.6% of the respondents reported that they had been exposed to violence in the last 10 months. Among the reported events, the most common form was verbal violence at 70.7%. The most common perpetrators of these cases were attendants of the patients in 87% of the cases. As an impact of these cases, 87.7% of the respondents felt emotional distress due to violent incidents. In addition, 6.3% reported physical injury, and 6.0% considered leaving their jobs. A statistically significant difference in the frequency of exposure to violence has been detected between age, gender, seniority, and province. The major contributing factors towards violence included lack of communication and lack of facilities along with lack of patient care.

**Conclusion:** Violence and aggression are one of the major issues of healthcare workplaces in Pakistan. Therefore, there is a dire need to improve working conditions, build effective communication, and improve security standards to create safe workplaces for healthcare professionals

**What We Already Know:**

❖ Workplace violence in healthcare is a recognized global concern, with healthcare workers being at a higher risk compared to other professions.
❖ Verbal abuse is the most common form of workplace violence encountered by doctors and nurses, particularly in developing countries like Pakistan.
❖ Factor contributing to violence in hospitals include lack of communication, inadequate facilities, and unrealistic patient expectations.
❖ Previous studies in Pakistan have generally been limited in scope—focused on specific cities, hospitals, or types of healthcare professionals—resulting in incomplete national data.

**What This Article Adds:**

❖ This study is among the first to provide comprehensive nationwide data on workplace violence against doctors in both public and private healthcare settings in Pakistan, including previously underrepresented region like Baluchistan and Gilgit Baltistan.
❖ It identifies specific demographic risk factors (e.g., younger age, early career stage, region, and shift timing) associated with higher incidences of violence.
❖ The research uncovers that a majority of perpetrators are patient attendants, emphasizing the need for targeted awareness and security interventions.
❖ It highlights that 67.8% of doctors feel unsafe at work, with 87.7% of doctor reporting distress and 6% considering leaving job, offering critical evidence for policy changes and safety protocol implementation across healthcare institutions.

## Introduction

The World Health Organization defines violence as the deliberate application of physical force or power, whether threatened or actual, directed at oneself, another person, or a group or community, which leads to, or has a strong potential to cause, injury, death, psychological harm, developmental issues, or deprivation [1]. Aggression is any behavior, including verbal threats, which involves attacking another person, animal, or object with the intent of harming the target [2]. Collectively, violence and aggression refer to behaviors that can result in harm to others. These behaviors may be expressed through actions or words, but the intent and physical harm inflicted are evident [3]. The necessity for guidelines addressing the short-term management of violence and aggression in healthcare, and community settings arises from their frequent occurrence and serious impact. Managing these behaviors are complex, influenced by both internal factors (such as personality traits, emotional distress, and anger management issues) and external factors (including the environment, social context, attitudes of aggressors, victim characteristics etc.) [4]. The focus of these guidelines is to ensure the safety of both staff and service users by offering methods to prevent and handle violent situations effectively. They are framed for healthcare professionals and individuals with mental health conditions, whether adults, adolescents, or children as well as their families and caregivers who rely on these services [5].

### Workplace violence in healthcare: A Global perspective

Globally, 23% of workers have encountered one of three types of violence and harassment including psychological, physical, or sexual. This includes 23% of women and 22% of men. Specifically, 18% of workers have faced psychological violence and harassment, while 9% have experienced physical violence, and 6% have encountered sexual harassment [6]. Although workplace violence impacts nearly all sectors, it is especially prevalent in healthcare settings, raising serious concerns for public health and occupational safety. Workers in healthcare and social services face the highest rates of violence-related injuries, being five times more likely to be injured compared to those in other industries. Furthermore, incidents of violence in the health sector are estimated to represent approximately 25% of all workplace violence occurrences [7,8]. Eighty-five percent of workplace violence incidents were categorized as Type II Customer/Client Workplace Violence, where employees are attacked by customers, clients, patients, students, inmates, or others receiving services. The overall prevalence of workplace violence against healthcare professionals was found to be 62.4%. Verbal abuse was the most common form, accounting for 61.2%, followed by psychological violence at 50.8%, threats at 39.5%, physical violence at 13.7%, and sexual harassment at 6.3% [11]. In 2018, healthcare professionals represented 73% of all nonfatal workplace injuries and illnesses related to violence. According to the World Health Organization (WHO), an estimated 8% to 38% of health workers encounter physical violence during their careers [8]. WHO acknowledges violence as a workplace risk, with a concerning 62% of healthcare workers reportedly affected by workplace violence (WPV) [12]. Furthermore, many are also subjected to or threatened by verbal aggression [9]. The assault rate against healthcare workers, as per the Healthcare Crime Survey conducted by the International Association for Healthcare Security and Safety Foundation (IAHSS Foundation), increased from 9.3 incidents per 100 beds in 2016 to 11.7 in 2018, marking the highest rate recorded since 2012 [10]. Although some organizations may have an established formal incident reporting system, numerous incidents particularly those involving bullying, verbal abuse, and harassment remain unreported due to multiple reasons including fear of retaliation, stigma, shame or in some cases insufficient awareness.

### Pakistani Context: Gaps and Urgency

The figures for a developing country like Pakistan are particularly troubling. A report from the Pakistan Medical Association stated that between 1995 and 2015, 128 doctors in Pakistan were killed due to violence [13]. Verbal abuse is the most common mistreatment faced by doctors, affecting 85% that means 85 out of 100 doctors experience this abuse, adversely impacting their mental health. Around 62% report moderate incidents like threats, while 38.1% encounter serious events [14]. 73.1% of nurses reported experiencing violence in the last year, with 53.4% facing physical violence, 57.3% verbal abuse, and 26.9% sexual violence. The main offenders were identified as male coworkers, patients, and attendants [15]. Another study reported a 10% prevalence of sexual harassment mainly nurses, with similar rates across government and private healthcare facilities. Patients’ relatives (47.8%) and staff members (32.6%) were the most common perpetrators [16].

#### Study Aim

Workplace violence (WPV) in healthcare significantly affects healthcare professionals, patients, and system performance. While provincial-level research on WPV exists, there is a lack of comprehensive national data in Pakistan. This study aims to fill that gap by collecting data from both government and private hospitals across the country. The primary objectives are to assess the prevalence of violence against doctors in the last 10 months, analyze the impact of gender, age, and work experience on the frequency of violence, and identify key risk factors. The findings will offer essential insights to help develop strategies for enhancing safety and reducing WPV in healthcare institutions nationwide.

## Methodology

### Study Design

A cross-sectional survey was conducted to examine the prevalence, nature of violence and aggression directed towards doctors across Pakistan.

### Sampling and Participant Selection

A multi-stage random sampling approach was applied to achieve a representative sample:

Using a random number generator, districts were chosen from each of Pakistan’s five provinces (Sindh, Punjab, Baluchistan, Khyber Pakhtunkhwa, and Gilgit-Baltistan) to ensure national geographic representation. In addition, the Capital Territory (Islamabad) and Azad Jammu and Kashmir (AJK) were considered as single entities.

In each selected district, two to three hospitals were chosen randomly, also through a random number generator, to capture diversity in healthcare settings.

Doctors with at least ten months of clinical experience were selected randomly within each hospital, a variety of departments covering morning, afternoon, and night shifts to capture the experiences of doctors working in different hours. Participant demographics including age, gender, province, job, experience and specialty were collected to allow for a comprehensive analysis of responses across different subgroups.

### Sample Size

The sample size was approximately 1,064 doctors, selected to provide a robust dataset for analysis, with an estimated confidence level of 95% and a margin of error around 3%. This size was chosen based on preliminary prevalence rates of workplace violence observed in similar studies.

### Survey Instrument

The survey used a structured questionnaire, which was administered in person by a co-author. It contained both closed-ended and Likert-scale questions to capture the frequency and type of aggression (physical and verbal) experienced by doctors. Definitions like (violence: Intentionally harm someone physically or emotionally) and (Aggression: action or behavior that is intended to harm someone’s emotions or well-being) were provided within the survey to ensure consistency in interpretation by all participants.

### Pilot Testing and Refinement

Pilot testing was conducted at Mardan Medical Complex to evaluate the clarity and reliability of the questionnaire. Based on feedback, two questions were removed to improve focus and readability. The final questionnaire was positively received with feedback, suggesting that it effectively captured relevant information.

### Ethics and Consent

Ethical approval is obtained from ethical committee of Bacha Khan medical college (ethical No 538/BKMC on 3^rd^ June 2024) before starting of data collection. This ethical approval is used in different site for data collection. Written informed consent was taken before filling of questionnaire and consent form is with questionnaire google form. Participation has to agree written consent before filling questionnaire. Consent form is attached in questionnaire form

### Data Collection Process

Data was collected after receiving ethical approval from the institutional administration, with written informed consent obtained from each participant before questionnaire filling. Each hospital was visited, and the questionnaire was personally distributed to doctors, allowing them to complete it. This approach ensured the confidentiality of the responses. The questionnaire was filled by 1003 doctors who agreed to take part in study by filling written informed consent. Participant were prospectively recruited from date 4 July 2024 to 20 December 2024 across hospital of five province in Pakistan. There is no retrospective data in this study

### Data Analysis

Data from completely filled questionnaires were entered and analyzed using an SPSS, v27. Statistical methods included descriptive analyses to quantify the prevalence of different types of aggression and inferential tests (e.g., chi-square, Logistic regression) to examine associations between demographic factors (such as age, gender, specialty, or shift) and experiences of violence.

## Results

### Demographic characteristics and prevalence of violent events

Table 1 contains the demographic characteristics (including age, gender, province, job title and clinical experience) of the study participants. A total of 1003 doctors participated in the study with a response rate of 94% (total 1068) with a nearly equal gender distribution i.e., 48.6% males and 51.4% females. The number of participants in the study where highest from KPK followed by Punjab, Sindh and Baluchistan. Prevalence of violence were found 77.6% across the country, while 22.4% of participants didn’t experience any type of violence or aggression in last 10 months.

**Table 1:**
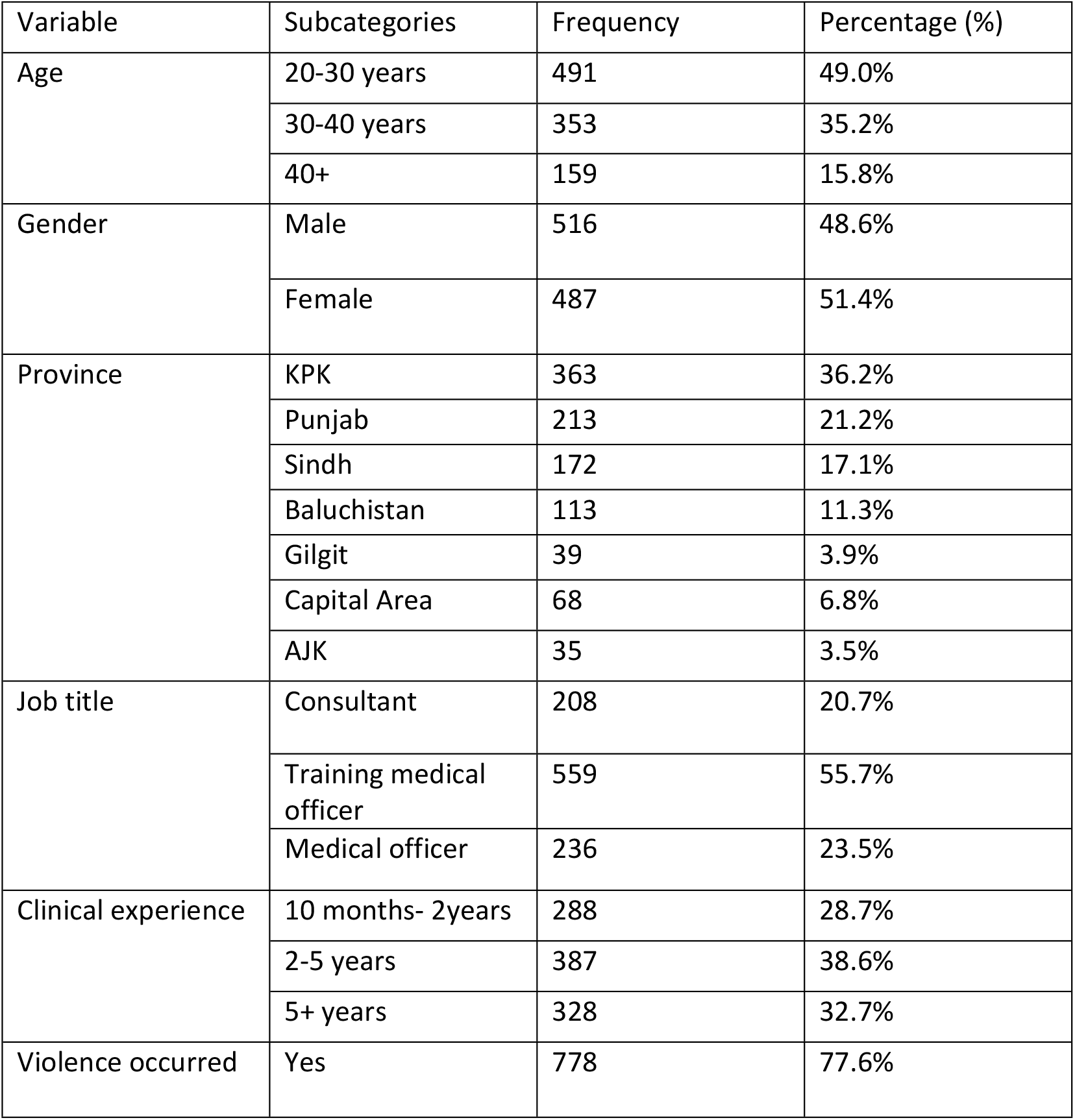
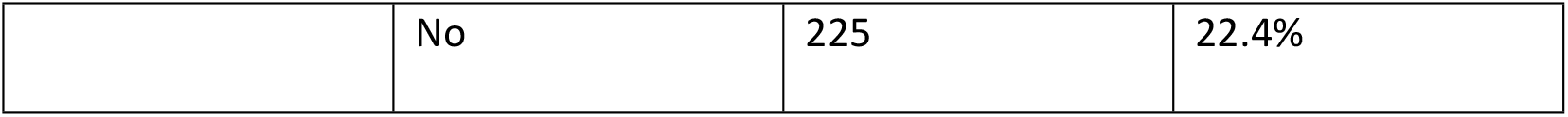
Demographic characteristics of the study population and prevalence of workplace violence.

While table 2 shows relation of demographic factors and workplace violence. In which, gender, experience, specialty and geographical location were found to have significant relation with violence (p value <.05).

**Table 2.** Demographic factors and workplace violence.

| Variable | Exposure to workplace violence |  | P-value | (X<sup>2</sup>)df |
| --- | --- | --- | --- | --- |
|  | Group A= Yes | Group B= No |  |  |
| Age (20-30) | 372 (47.8%) | 119 (52.8%) | .407 | (1.789<sup>a</sup>)2 |
| Gender (male) | 413 (53.08%) | 103 (45.8%) | .053 | (3.731<sup>a</sup>)1 |
| Province (KPK) | 265 (34.0%) | 98 (43.6%) | .000 | (40.389<sup>a</sup>)6 |
| Job title<br>(training<br>medical officer) | 426 (54.8%) | 133 (59.1%) | .501 | (1.384<sup>a</sup>)2 |
| Specialty<br>(general<br>medicine) | 155 (19.9%) | 38 (16.8%) | .029 | (32.286<sup>a</sup>)19 |
| Experience (2-5 years) | 324 (41.6%) | 63 (28.0%) | .000 | (32.282<sup>a</sup>)2 |

Figure 1 indicates the geographical distribution of the respondents. The study also revealed doctors from Khyber Pakhtunkhwa (KPK) were more likely to experience workplace violence as compared to other regions (p<0.001). Doctors in their early career (p<0.001) and from general medicine specialty (p=0.029) reported higher incidences of workplace violence.

**Figure 1.**
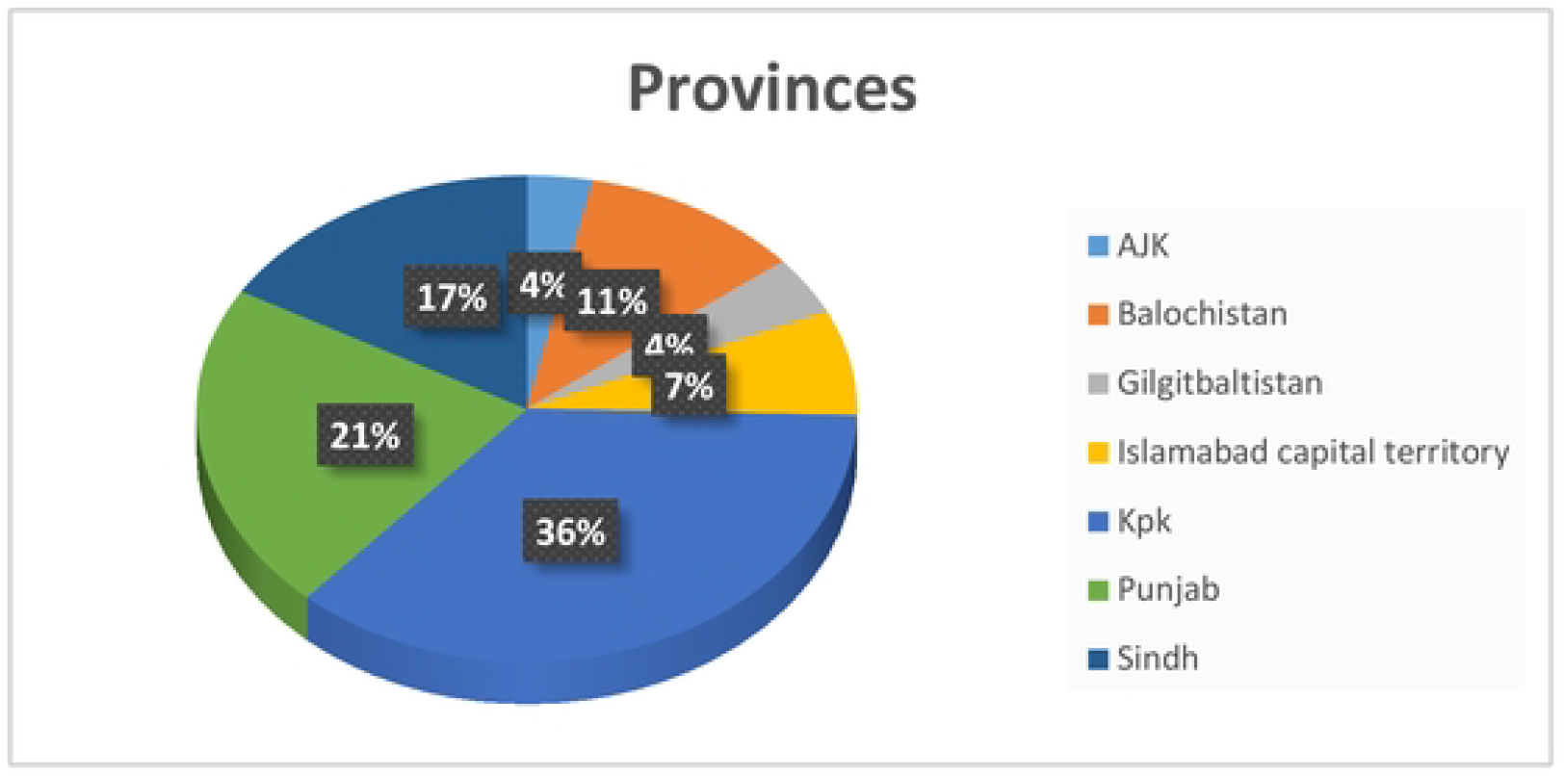
The distribution of the participants as per province.

**Figure 2.**
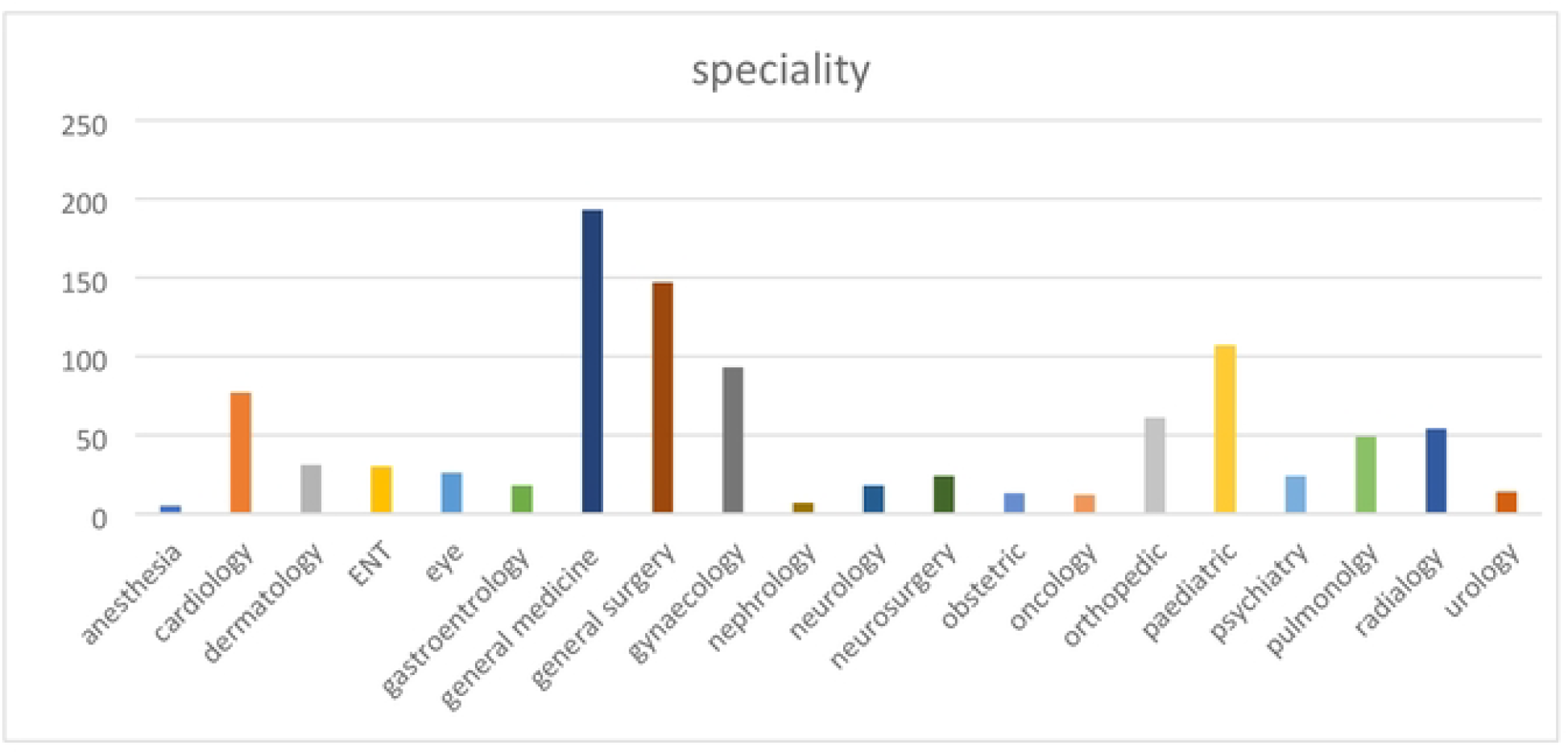
shows the distribution of the participants as per specialty

### Frequency, Perpetrators and impact on doctors

The workplace violence over the last 10 months was reported by 77.6% of the doctors (Table 1). Among them (n=778), the majority of the participants 56.6% reported both experiencing and witnessing the violent event, and the most common type of violence was verbal violence (70.7%) figure 3.

**Figure 3.**
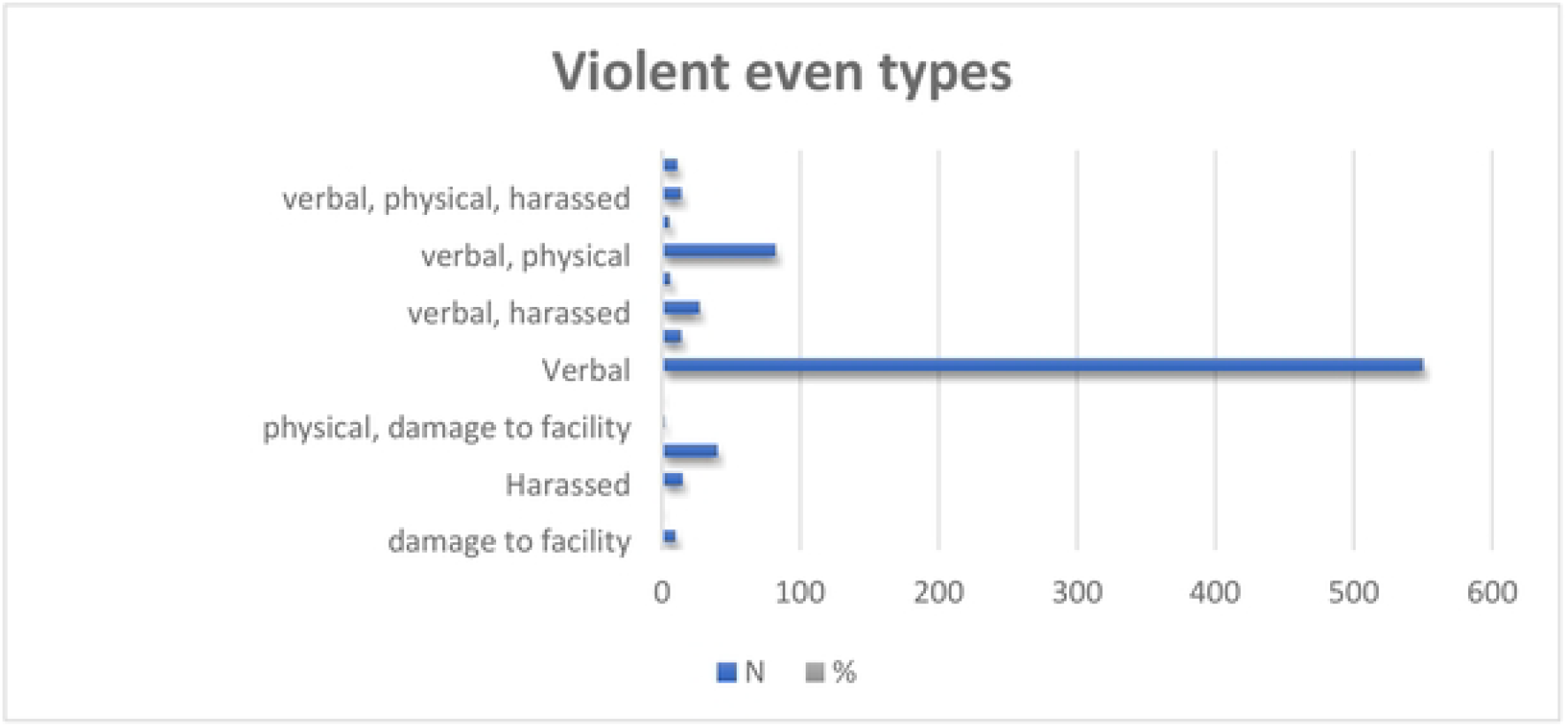
violent even types

Most commonly violent events were reported in tertiary care settings (78.1%) indicating that larger hospitals, where patient loads are presumably higher, are at a higher risk of experiencing violent events of overcrowding, long waiting hours, and patient dissatisfaction.

In most cases (87%), attendants were involved followed by patients (10.9%). The emotional and psychological effect of violence on healthcare workers was evident, as 87.7% of the respondents said that they experienced emotional distress, while 6% considered leaving their job. (table 3)

**Table 3.** Details of the workplace violence against doctors.

| Variable | Subcategories | Frequency | Percentage |
| --- | --- | --- | --- |
| Violence witnessed or experienced | Experienced | 175 | 22.5% |
|  | Witnessed | 163 | 21.0% |
|  | Both | 440 | 56.6% |
| Frequency of violent events | 1 | 170 | 21.9% |
|  | 2-3 | 276 | 35.5% |
|  | 4-5 | 158 | 20.3% |
|  | 6+ | 174 | 22.4% |
| Healthcare setting | Primary care hospital | 14 | 1.8% |
|  | Secondary care hospital | 156 | 20.1% |
|  | Tertiary care hospital | 608 | 78.1% |
| Perpetrator | Attendant | 677 | 87.0% |
|  | Patient | 85 | 10.9% |
|  | Nurse | 9 | 1.2% |
|  | Security staff | 7 | 0.9% |
| Shift | Morning shift | 268 | 34.4% |
|  | Evening shift | 221 | 28.4% |
|  | Night shift | 289 | 37.1 |
| Reason of violence | Lack of communication | 327 | 42.0% |
|  | Lack of facility | 374 | 48.1% |
|  | Lack of patient care | 77 | 9.9% |

### Contributing Factors, Recommendations, and Workplace Security Perceptions Among Doctors

The study participants reported lack of awareness, workload on doctors, lack of facilities, and higher patient expectations as the most frequent contributing factors. Most of the doctors recommended improvements in healthcare services and public awareness campaigns as the solutions. Doctors’ perception of workplace security highlighted widespread concerns, with 67.8% reporting feeling unsafe at work. When given a choice 87.5% of the respondents preferred medium pay with job security over high pay with less security indicating that workplace safety is a major concern in career decision-making. (Table 4)

**Table 4:** Contributing Factors, Recommendations, and Workplace Security Perceptions Among Doctors.

| Variables | Categories | Frequency | Percentage |
| --- | --- | --- | --- |
| Contributing factors | High expectations | 103 | 10.3% |
|  | Lack of awareness | 448 | 44.7% |
|  | Lack of facility | 128 | 12.8% |
|  | Shortage of healthcare staff | 74 | 7.4% |
|  | Workload on doctors | 250 | 24.9% |
| Recommendations | Improvement in healthcare services | 451 | 45.0% |
|  | Increase healthcare staff | 70 | 7.0% |
|  | Overall public awareness | 330 | 32.9% |
|  | Provision of security | 152 | 15.2% |
| What to choose | High pay with less security | 143 | 14.3% |
|  | Medium pay with less security | 860 | 85.7% |
| Feel secure at work | Yes | 323 | 32.2% |
|  | No | 680 | 67.8% |

## Discussion

### Key findings

This research points to the widespread violence against doctors in healthcare across Pakistan, with a particularly high prevalence of verbal abuse [70.7%]. The previous studies also show a high prevalence of violence. Verbal violence was the most commonly experienced form (33.9%), followed by physical violence (6.6%) [21]. Verbal. Violence was more prevalent compared to physical violence [20]. Several factors contribute to this violence which includes high expectations from doctors, inadequate facilities, and excessive workloads. Despite the high rates of violence, most doctors still feel insecure [67.8%] in their workplaces and prioritize job security over higher pay [85.7%]. Workplace violence may lead to lower job satisfaction and increased turnover among healthcare professionals [25]. These results highlight the fact that better healthcare infrastructure, along with enhanced security and public awareness campaigns, should be initiated to reduce such occurrences. Furthermore, there is a dire need of proper emotional care, to be given to affected doctors.

### Comparison with previous literature

Violence and aggression against healthcare professionals, specifically doctors are a major growing concern globally.as it has some serious consequences directly on doctor’s mental health which gives indirect effects on their work efficiency and patient’s care. Recently this concern is growing much rapidly in our country (Pakistan). According to a study in 2018 in Karachi, one-third of the participants (healthcare workers) had experienced some form of violence in the last 12 months [17]. According to another study across the country in Pakistan in 2020, more than one-third (38.4%) healthcare workers reported having experienced any form of violence in the last 6 months [18]. However, the literature on the assessment of workplace violence against doctors is still limited, and there is a lot of space for investigating this rapidly growing concern.

The issue of violence against doctors in Pakistan remains understudied with several key gaps in the literature. Most of the studies concentrate on immediate causes and cross-sectional data, leaving out the long-term psychological impact on doctors and the role of gender, regional, and sectoral differences in prevalence of violence. Workplace violence against healthcare workers is a widespread phenomenon with very severe consequences for the individuals affected and their organizations [28]. Moreover, socio-cultural, economic, and political factors contributing to such violence have not been sufficiently explored. Also the interventions and legal frameworks are not studied in relation to their effectiveness. De-escalation of violence training was effective in improving the confidence of healthcare providers in coping with patient aggression [23].

Workplace violence against doctors in public teaching hospitals is significantly associated with environmental, patient-related, and systemic factors [24]. The understanding and the more effective policies in the protection of healthcare professionals would be enhanced if these gaps were addressed, such as the integration of conflict management into medical training. In one study the review of the evidence, discussions with experts, and pilot findings highlight the potential value of incorporating comprehensive conflict management training in medical education [26]. Another study results suggest that the educational intervention was effective in improving medical students’ attitudes toward conflict management [27].

This study on violence against doctors in Pakistan partially addresses some of the important key gaps in the literature but also leaves several areas underexplored. The study provides empirical evidence on factors such as age, gender, job title, and clinical experience, and highlights that clinical experience is a significant predictor of violent incidents.

Similar to our study, which shown high prevalence of violence among those who have experience less than five years, another study mentioned that professionals who had less than five years of experience and worked in shifts, especially evening shifts, experience violence more frequently [22]. Another study shows that patient-led episodes of verbal violence are more prevalent in Asian countries, especially in the emergency department, psychiatric wards, and intensive care units, mostly faced by junior doctors and residents [19].

### Strength and Limitation

On the contrary, though study emphasis was focused on clinical experience, there lacks in-depth considerations on other areas such as efficiency of interventions offered, legal guarantees or training offered for conflict resolutions that remain among other key potential for future research subjects. Therefore, there are a few limitations that should be considered like it includes self-reported data, along with that there is also some elements of recall bias. Also, the study centers around doctors, which could limit its generalizability to other healthcare professionals, such as nurses or support staff, who may experience varied levels of violence

Despite the above-mentioned limitations, this research has bridged a critical information gap through providing empirical evidence regarding violence in healthcare settings in Pakistan. The present study regarding violence and aggression towards doctors in Pakistan throws light upon factors that trigger violent incidents within the healthcare sector. It also covers data from Baluchistan and Gilgit Baltistan which were previously not included. The findings can be used to influence policy changes and the implementation of more stringent safety measures, ultimately leading to a safer working environment for healthcare professionals. One of the strengths of this research is its comprehensive approach, which covers a large sample size of 1003 allowing an in-depth analysis of various demographic factors such as age, gender, job title, province, and clinical experience. Such findings are crucial for the establishment of specific intervention programs aimed at improving safety strategies for healthcare staff.

Suggestions for the future studies, and research gap in this study is that there is lack of qualitative data here. Along with that, this study has hospital type variability, culture and regional differences, which need to be address in future studies.

## Conclusion

This research work reflects intense violence rate in health care settings of Pakistan and points towards the immediate need for corrective action against the causative factors for violence. Improvement in working conditions, communication, and security can work well in the creation of a safer environment and would support the healthcare professionals better. Thus, it would contribute significantly for the overall improvement in quality of patient care.

## Data Availability

All relevant data are within the manuscript

## Notes

### Competing Interest Statement

The authors have declared no competing interest.

### Funding Statement

The author(s) received no specific funding for this work.

### Author Declarations

The study followed ethical guidelines as approved by the ethical committee of bacha khan medical college, ethical No 538/BKMC on 3rd June 2024. Participation was voluntary, with confidentiality assured for all the respondents.

## References

1. Rutherford A, Zwi AB, Grove NJ, Butchart A. Violence: a glossary. J Epidemiol Community Health. 2007;61(8):676–680. doi:10.1136/jech.2005.043711

2. Soreff SM, Wadhwa R, Arif H. Aggression. In: StatPearls. Treasure Island (FL): StatPearls Publishing; November 13, 2024.

3. Liu J. Concept analysis: aggression. Issues Ment Health Nurs. 2004;25(7):693–714. doi:10.1080/01612840490486755

4. Liu J, Ma H, He YL, et al. Mental health system in China: history, recent service reform and future challenges. World Psychiatry. 2011;10(3):210–216. doi:10.1002/j.2051-5545.2011.tb00059.x

5. National Collaborating Centre for Mental Health (UK). Violence and Aggression: Short-Term Management in Mental Health, Health and Community Settings. London: British Psychological Society (UK); 2015.

6. National Institute for Health and Care Excellence. Violence and aggression: short-term management in mental health, health and community settings | Guidance | NICE. Nice.org.uk. Published May 28, 2015. https://www.nice.org.uk/guidance/ng10

7. Crabtree S. Global Study: 23% of Workers Experience Violence, Harassment. Gallup.com. Published December 14, 2022. https://news.gallup.com/opinion/gallup/406793/global-study-workers-experience-violence-harassment.aspx

8. U.S. Bureau of Labor Statistics. Workplace Violence in Healthcare, 2018. https://www.bls.gov. Published April 2020. https://www.bls.gov/iif/oshwc/cfoi/workplace-violence-healthcare-2018.htm

9. Di Martino V. Workplace violence in the health sector: country case studies Brazil, Bulgaria, Lebanon, Portugal, South Africa, Thailand, and an additional Australian study. Geneva: International Labour Organization; 2002:3–42.

10. World Health Organization. Preventing violence against health workers. https://www.who.int. Published 2022. https://www.who.int/activities/preventing-violence-against-health-workers

11. Hoffman R. Violence Against Healthcare Workers. Patient Safety. 2019;1(2):2–2. https://patientsafetyj.com/index.php/patientsaf/article/view/reducing-violence

12. Rock A. Statistics You Should Know: Workplace Violence in Healthcare. Campus Safety Magazine. Published July 3, 2019. Accessed November 12, 2019. https://www.campussafetymagazine.com/hospital/workplace-violence-in-healthcare-statistics/

13. Liu J, Gan Y, Jiang H, et al. Prevalence of workplace violence against healthcare workers: a systematic review and meta-analysis. Occup Environ Med. 2019;76(12):927–937. doi:10.1136/oemed-2019-105849

14. Beech B, Leather P. Workplace violence in the health care sector: A review of staff training and integration of training evaluation models. Aggression and Violent Behavior. 2006;11(1):27–43. doi:10.1016/j.avb.2005.05.004

15. Ahmed F, Khizar Memon M, Memon S. Violence against doctors, a serious concern for healthcare organizations to ponder about. Ann Med Surg (Lond). 2017;25:3–5. Published 2017 Nov 15. doi:10.1016/j.amsu.2017.11.003

16. Jafree SR. Workplace violence against women nurses working in two public sector hospitals of Lahore, Pakistan. Nurs Outlook. 2017;65(4):420–427. doi:10.1016/j.outlook.2017.01.008

17. Somani R, Karmaliani R, Mc Farlane J, Asad N, Hirani S. Sexual Harassment towards Nurses in Pakistan: Are we Safe? International Journal of Nursing Education. 2015;7(2):286. doi:10.5958/0974-9357.2015.00120.8

18. Baig LA, Shaikh S, Polkowski M, et al. Violence Against Health Care Providers: A Mixed-Methods Study from Karachi, Pakistan. J Emerg Med. 2018;54(4):558-566.e2. doi:10.1016/j.jemermed.2017.12.047

19. Shaikh S, Baig LA, Hashmi I, et al. The magnitude and determinants of violence against healthcare workers in Pakistan. BMJ Glob Health. 2020;5(4):e002112. Published 2020 Apr 15. doi:10.1136/bmjgh-2019-002112

20. Kumari A, Kaur T, Ranjan P, Chopra S, Sarkar S, Baitha U. Workplace violence against doctors: Characteristics, risk factors, and mitigation strategies. J Postgrad Med. 2020;66(3):149–154. doi:10.4103/jpgm.JPGM_96_20

21. Kumari A, Ranjan P, Sarkar S, Chopra S, Kaur T, Baitha U. Identifying Predictors of Workplace Violence Against Healthcare Professionals: A Systematic Review. Indian J Occup Environ Med. 2022;26(4):207–224. doi:10.4103/ijoem.ijoem_164_21

22. Baig L, Tanzil S, Shaikh S, Hashmi I, Khan MA, Polkowski M. Effectiveness of training on de-escalation of violence and management of aggressive behavior faced by health care providers in a public sector hospital of Karachi. Pak J Med Sci. 2018;34(2):294–299. doi:10.12669/pjms.342.14432

23. Yousaf MR, Imran H, Nadeem H, Jamil H, Butt MA, Khan RM. Workplace violence against doctors in public teaching hospitals in Pakistan: a cross-sectional study evaluating the contributing factors. J Soc Prev Adv Res KEMU. 2025;3(4):14–19. doi:10.21649/jspark.v3i4.715.

24. Cochran N, Charlton P, Reed V, Thurber P, Fisher E. Beyond fight or flight: The need for conflict management training in medical education. Conflict Resolution Quarterly. 2018;35(4):393–402. doi:10.1002/crq.21218

25. Mohseni F, Mohammadi A, Mafinejad MK, Khajavirad N, Basiri K, Gruppen LD. Teaching conflict management to medical students: a randomized controlled trial. BMC Med Educ. 2024;24(1):1507. Published 2024 Dec 20. doi:10.1186/s12909-024-06514-8

26. Witteman HO, Haverfield J, Tannenbaum C. COVID-19 gender policy changes support female scientists and improve research quality. Proc Natl Acad Sci U S A. 2021;118(6):e2023476118. doi:10.1073/pnas.2023476118

27. Balducci C, Rafanelli C, Menghini L, Consiglio C. The Relationship between Patients’ Demands and Workplace Violence among Healthcare Workers: A Multilevel Look Focusing on the Moderating Role of Psychosocial Working Conditions. Int J Environ Res Public Health. 2024;21(2):178. Published 2024 Feb 4. doi:10.3390/ijerph21020178

28. Vento S, Cainelli F, Vallone A. Violence Against Healthcare Workers: A Worldwide Phenomenon With Serious Consequences. Front Public Health. 2020;8:570459. Published 2020 Sep 18. doi:10.3389/fpubh.2020.570459

